# Antibody therapy reverses biological signatures of COVID-19 progression

**DOI:** 10.1101/2021.12.21.21268197

**Authors:** M. Cyrus Maher, Leah B. Soriaga, Anil Gupta, Julia di Iulio, Sarah Ledoux, Megan J. Smithey, Andrea L. Cathcart, Kathleen McKusick, David Sun, Melissa Aldinger, Elizabeth Alexander, Lisa Purcell, Xiao Ding, Amanda Peppercorn, Daren Austin, Erik Mogalian, Wendy W. Yeh, Adrienne E. Shapiro, Davide Corti, Herbert W. Virgin, Phillip S. Pang, Amalio Telenti

## Abstract

Understanding who is at risk of progression to severe COVID-19 is key to effective treatment. We studied correlates of disease severity in the COMET-ICE clinical trial that randomized 1:1 to placebo or to sotrovimab, a monoclonal antibody for the treatment of SARS-CoV-2 infection. Several laboratory parameters identified study participants at greater risk of severe disease, including a high neutrophil-lymphocyte ratio (NLR), a negative SARS-CoV-2 serologic test and whole blood transcriptome profiles. Sotrovimab treatment in these groups was associated with normalization of NLR and the transcriptomic profile, and with a decrease of viral RNA in nasopharyngeal samples. Transcriptomics provided the most sensitive detection of participants who would go on to be hospitalized or die. To facilitate timely measurement, we identified a 10-gene signature with similar predictive accuracy. In summary, we identified markers of risk for disease progression and demonstrated that normalization of these parameters occurs with antibody treatment of established infection.

## Introduction

Sotrovimab is a human monoclonal antibody (mAb) derived from an antibody isolated from a person recovered from SARS-CoV infection. This mAb broadly neutralizes SARS-CoV-2, SARS-CoV and other related animal sarbecoviruses^1-3^. Sotrovimab targets a highly conserved epitope in the SARS-CoV-2 Spike protein located in a site outside the receptor-binding motif (RBM), thus retaining in vitro activity against current SARS-CoV-2 variants of concern (VOC) (Alpha, Beta, Gamma, Delta, Omicron) and variants of interest (VOI)^4,5^. The majority of mAbs developed for COVID-19 bind to the RBM, which engages the angiotensin-converting enzyme 2 (ACE2) receptor. The RBM is one of the most mutable and immunogenic regions of the virus^6^. As a result, RBM antibodies as single agents and even when used in combination have not retained activity against some VOC/VOI^7-10^, particularly against Omicron VOC ^5,11,12^.

Sotrovimab was recently tested in a multicenter, double-blind, phase 3 clinical trial (COMET-ICE, ClinicalTrials.gov NCT04545060) that recruited non-hospitalized participants with symptomatic COVID-19, and at least one known risk factor (age and/or comorbidities) for severe disease progression. Participants were randomized to a single intravenous infusion of sotrovimab 500 mg or placebo. In the interim analysis of the trial, sotrovimab significantly reduced the risk of all-cause hospitalization (>24 hours) or death from COVID-19^1^. The final data, now presented in preprint^2^, shows that among 1057 participants randomized (sotrovimab, 528; placebo, 529), all-cause hospitalization longer than 24 hours or death was significantly reduced with sotrovimab (6/528 [1%]) vs placebo (30/529 [6%]) by 79% (95% CI, 50% to 91%; p<0.001).

While the impact of sotrovimab was profound, the relatively low rate of hospitalization or death amongst participants considered at risk for poor disease outcomes in the placebo arm led us to investigate if additional biomarkers or biomarker profiles beyond the known demographic and comorbid conditions could be identified. The setting of a randomized, controlled clinical trial presented a unique opportunity to identify signals of disease progression that resolved in response to treatment and could thus be used both to provide insights into COVID-19 pathogenesis, and as potential surrogate endpoints in the design of future trials. Thus, the present study aimed at identifying baseline correlates of hospitalization and severe disease/death in an at-risk population based on routine laboratory parameters, SARS-CoV-2 serology and on transcriptome analysis. It then sought to assess the impact of antibody treatment on these parameters. This approach enabled assessment of the impact of antibody treatment on populations with different intrinsic risks of disease progression, and the identification and testing of surrogates of treatment response.

## Results

### Identification of study participants at high risk of progression of COVID-19 using clinical laboratory values

COMET-ICE^2^ included 1057 adults with a positive local polymerase-chain-reaction or antigen SARS-CoV-2 test result and onset of symptoms within the prior 5 days (**Table 1**). We analyzed 63 available central laboratory parameters for their association to hospitalization or death (**Suppl. File 1**). On Day 1, pre-dose, white blood cell proportions were most predictive of eventual hospitalization or death (**Table 2**). White blood cell proportions were also quantified by the neutrophil to lymphocyte ratio (NLR; area under a receiver operating characteristic curve, AUC=0.81). We found that an NLR greater or equal 6 provided an optimal cutoff for the highest enrichment for disease progression and is hereafter defined as ‘high NLR’ (**Suppl. Fig. S1**). Of the 36 hospitalizations or deaths that occurred in the COMET-ICE study, 29 of these were observed among the 903 participants with available NLR at Day 1, the day of dosing. NLR had a sensitivity of 45% [28%-63%] and a specificity of 95% [93%-96%] for the prediction of all-cause hospitalization or death (Fisher’s exact p<0.001). NLR normalized more rapidly in participants receiving sotrovimab (**Figure 1**).

**Table 1.**
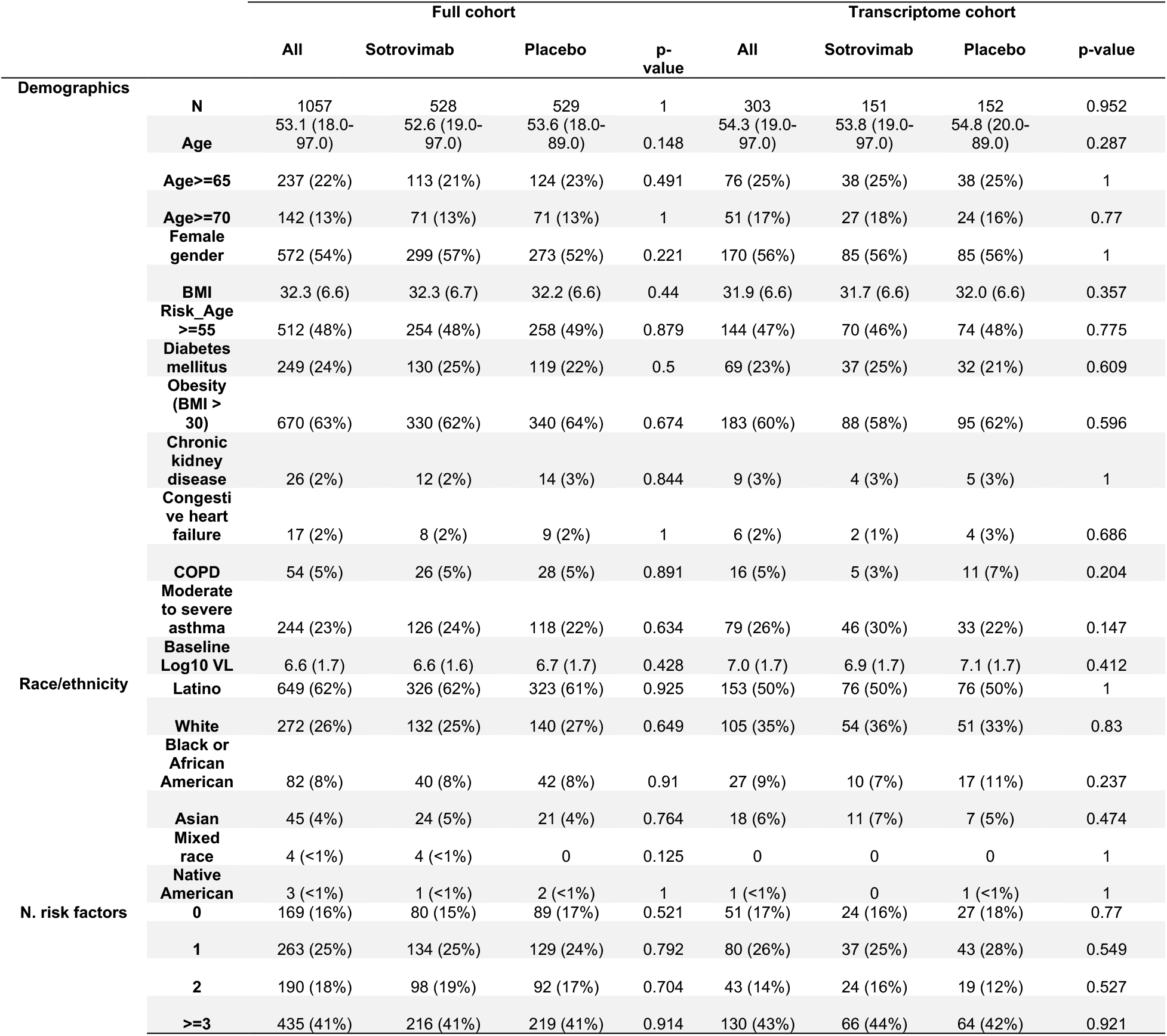
Baseline Demographic and Disease Characteristics. Numbers in parentheses represent the percent of all study participants in that category when denoted with “%”; otherwise, they represent the standard deviation. The numbers outside the parentheses are counts when represented with “%” in the parentheses. Otherwise, they are the mean value of that variable.

**Table 2.**
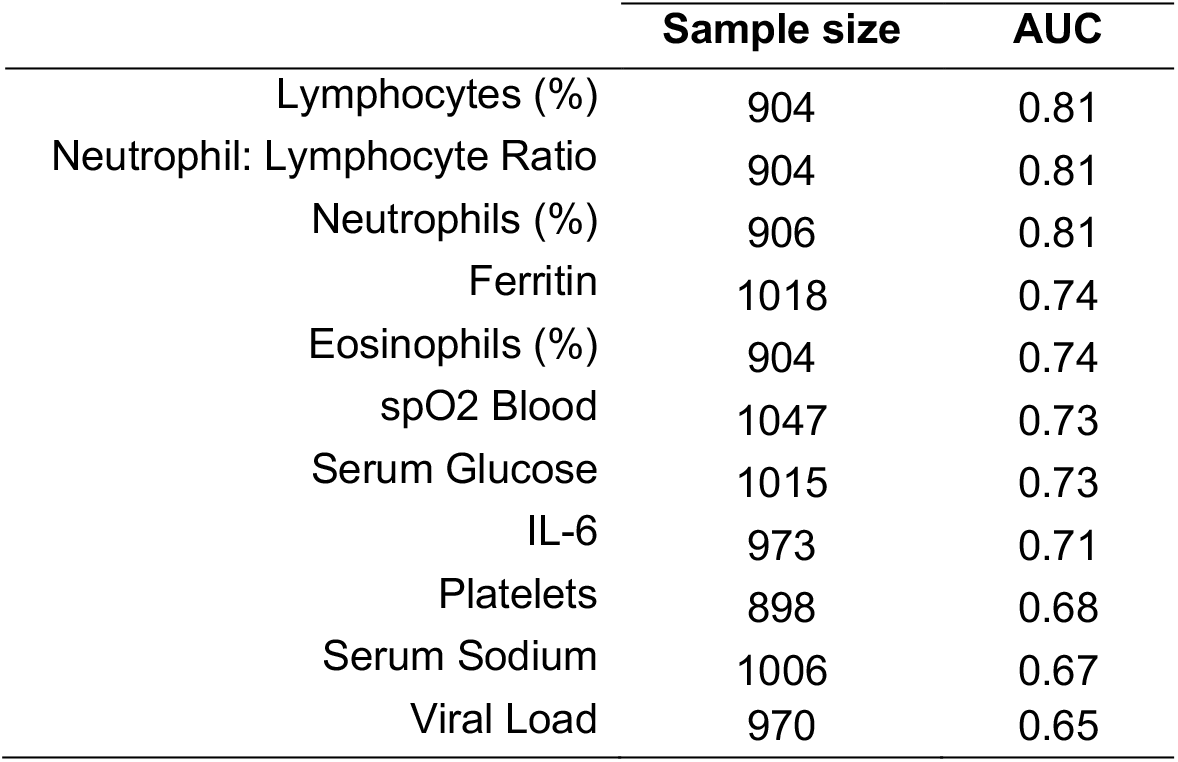
Top laboratory predictors of hospitalization at baseline. Area under the curve (AUC) and sample sizes for top clinical predictors of hospitalization or death based on baseline, Day 1 values. For a complete list of 63 parameters, see **Suppl. File 1**.

|  | **Sample size** | **AUC** |
| --- | --- | --- |
| Lymphocytes (%) | 904 | 0.81 |
| Neutrophil: Lymphocyte Ratio | 904 | 0.81 |
| Neutrophils (%) | 906 | 0.81 |
| Ferritin | 1018 | 0.74 |
| Eosinophils (%) | 904 | 0.74 |
| spO2 Blood | 1047 | 0.73 |
| Serum Glucose | 1015 | 0.73 |
| IL-6 | 973 | 0.71 |
| Platelets | 898 | 0.68 |
| Serum Sodium | 1006 | 0.67 |
| Viral Load | 970 | 0.65 |

**Figure 1.**
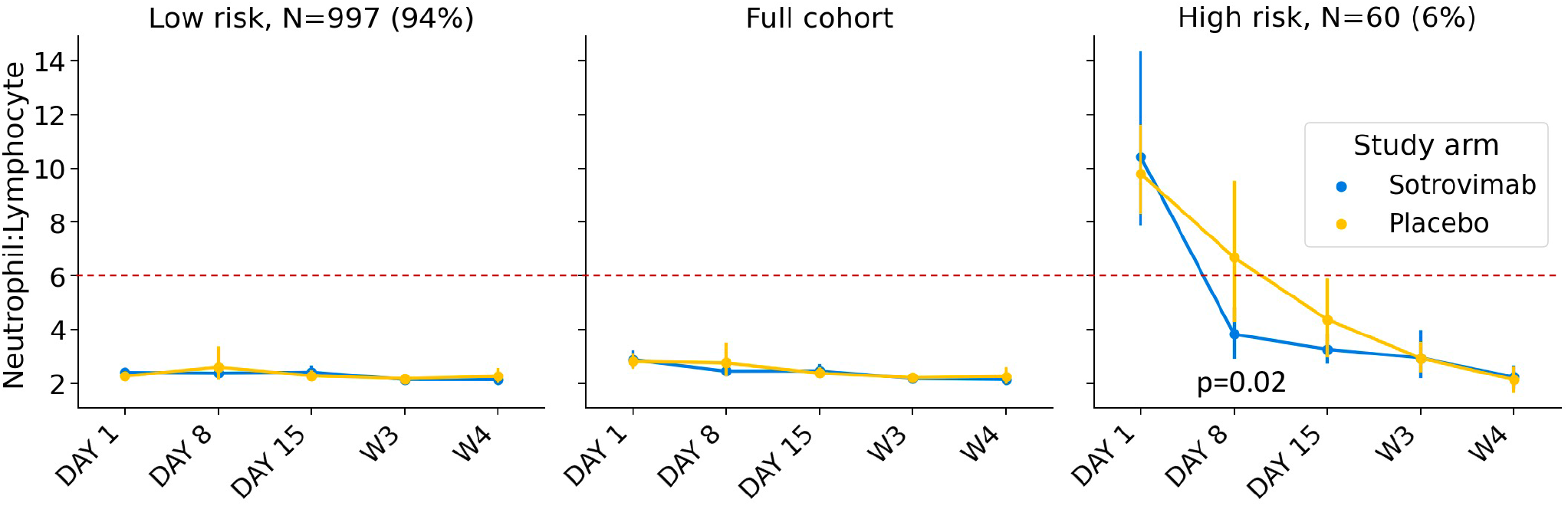
Response to sotrovimab in high-risk group defined by neutrophil lymphocyte ratio (NLR). The time trend of NLR for sotrovimab versus placebo treated patients (hue) in the full cohort and low risk and high-risk groups as defined by NLR ≥ 6. Error bars indicate the 95% confidence interval on the mean.

### The role of SARS-CoV-2 serostatus in defining risk of progression of COVID-19

Having SARS-CoV-2 anti-nucleocapsid antibodies provides protection against SARS-CoV-2 re-infection^13^. There is more limited information, however, on how serostatus may associate with severity of disease during acute infection. In the current study, seropositivity at baseline may indicate prior infection by SARS-CoV-2 or that a participant is already seroconverting during an acute infection episode. Seropositivity rates varied significantly by race/ethnicity, **Table 3**. Seropositivity was also associated with lower viral RNA in nasopharyngeal swabs at baseline: mean 4.2 vs 6.4 log10 viral RNA in seropositive versus seronegative participants (Mann-Whitney U p=1e-36); **Table 3**. Of the participants who received placebo and the serology results were available, 4/97 (4%) of seropositive participants progressed to hospitalization and/or death compared with 25/375 (7%) of the seronegative who progressed to hospitalization and/or death before Day 29. In participants who received placebo, there were no deaths or ICU admissions in those who were seropositive at baseline compared to 4 deaths (2 deaths before Day 29 and 2 additional deaths that occurred after Day 29) and 9 ICU admissions (2.4%) in those who were seronegative at baseline.

**Table 3.**
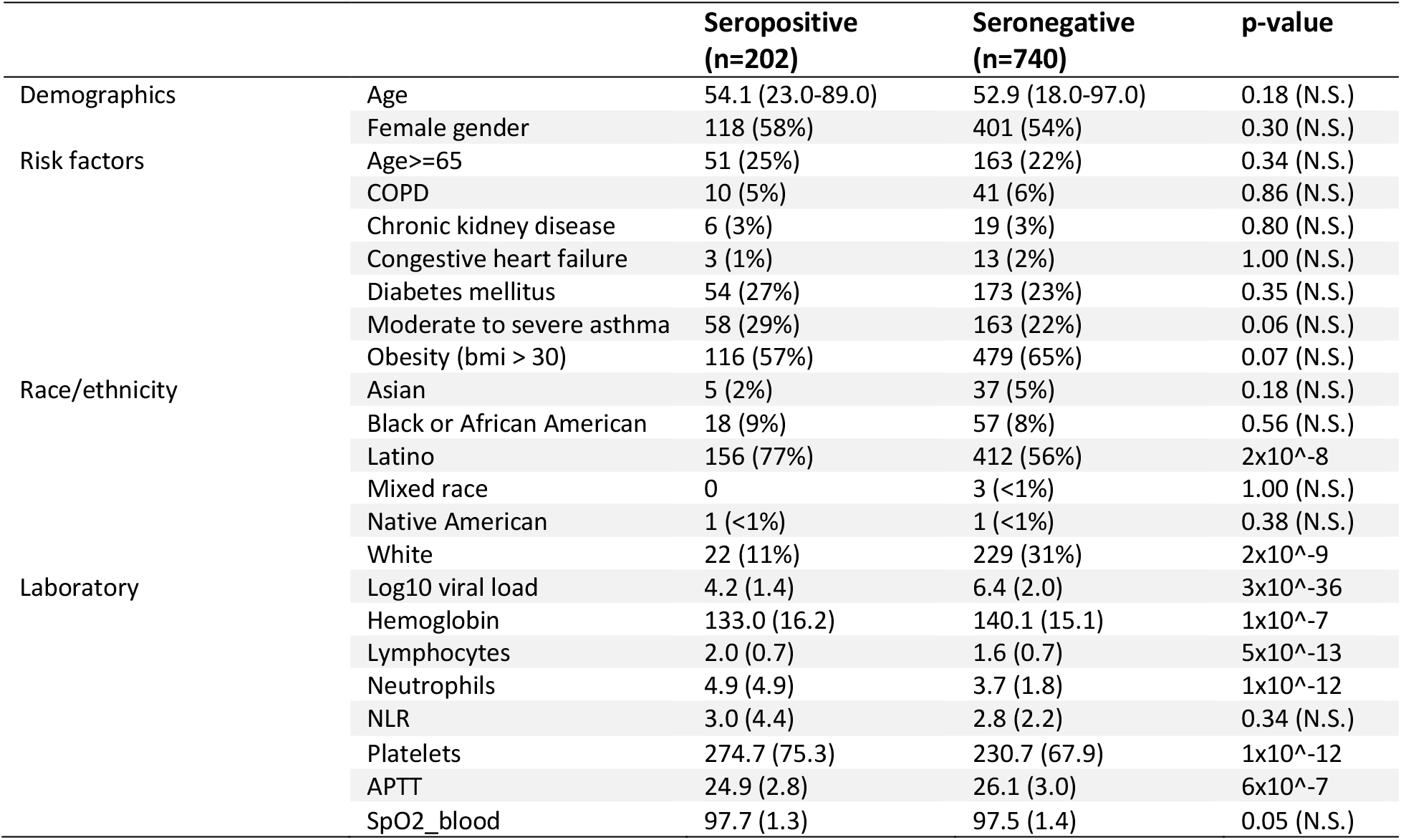
Comparison of representative baseline values between seropositive and seronegative patients. Numbers in parentheses represent the percent of all participants in that category when denoted with “%”; otherwise, they represent the standard deviation. The numbers outside the parentheses are counts when represented with “%” in the parentheses. Otherwise, they are the mean value of that variable. P-value significance thresholds are based on a Bonferroni correction (p=0.05 / 23 comparisons). P-values are calculated using either the Mann Whitney U or Fisher’s exact test depending on whether the variable is continuous or binary, respectively.

|  |  | **Seropositive**<br>**(n=202)** | **Seronegative**<br>**(n=740)** | **p-value** |
| --- | --- | --- | --- | --- |
| Demographics | Age | 54.1 (23.0-89.0) | 52.9 (18.0-97.0) | 0.18 (N.S.) |
|  | Female gender | 118 (58%) | 401 (54%) | 0.30 (N.S.) |
| Risk factors | Age≥65 | 51 (25%) | 163 (22%) | 0.34 (N.S.) |
|  | COPD | 10 (5%) | 41 (6%) | 0.86 (N.S.) |
|  | Chronic kidney disease | 6 (3%) | 19 (3%) | 0.80 (N.S.) |
|  | Congestive heart failure | 3 (1%) | 13 (2%) | 1.00 (N.S.) |
|  | Diabetes mellitus | 54 (27%) | 173 (23%) | 0.35 (N.S.) |
|  | Moderate to severe asthma | 58 (29%) | 163 (22%) | 0.06 (N.S.) |
|  | Obesity (bmi > 30) | 116 (57%) | 479 (65%) | 0.07 (N.S.) |
| Race/ethnicity | Asian | 5 (2%) | 37 (5%) | 0.18 (N.S.) |
|  | Black or African American | 18 (9%) | 57 (8%) | 0.56 (N.S.) |
| | Latino | 156 (77%) | 412 (56%) | $2 \times 10^{-8}$ |
|  | Mixed race | 0 | 3 (<1%) | 1.00 (N.S.) |
|  | Native American | 1 (<1%) | 1 (<1%) | 0.38 (N.S.) |
| | White | 22 (11%) | 229 (31%) | $2 \times 10^{-9}$ |
| Laboratory | Log10 viral load | 4.2 (1.4) | 6.4 (2.0) | $3 \times 10^{-36}$ |
| | Hemoglobin | 133.0 (16.2) | 140.1 (15.1) | $1 \times 10^{-7}$ |
| | Lymphocytes | 2.0 (0.7) | 1.6 (0.7) | $5 \times 10^{-13}$ |
| | Neutrophils | 4.9 (4.9) | 3.7 (1.8) | $1 \times 10^{-12}$ |
|  | NLR | 3.0 (4.4) | 2.8 (2.2) | 0.34 (N.S.) |
| | Platelets | 274.7 (75.3) | 230.7 (67.9) | $1 \times 10^{-12}$ |
| | APTT | 24.9 (2.8) | 26.1 (3.0) | $6 \times 10^{-7}$ |
|  | SpO2_blood | 97.7 (1.3) | 97.5 (1.4) | 0.05 (N.S.) |

Of the 202 seropositive participants at baseline, 6 (3%) were hospitalized or died: 4/96 (4.2%) received placebo and 2/106 (1.9%) received sotrovimab. Notably, as the COMET-ICE study captured all-cause hospitalizations or death, all 4 seropositive participants in the placebo arm were hospitalized with COVID-19 diagnoses, while the two seropositive participants in the sotrovimab arm were hospitalized with events potentially unrelated to COVID-19 (one instance of diabetic foot, and one instance of non-small cell lung cancer). No sotrovimab-treated participants died or were admitted to the ICU.

### Identification of high-risk cluster using transcriptomics

We used whole blood transcriptomics to define additional laboratory-based predictors of disease progression and response to treatment. In theory, such transcriptome signatures could provide complementary insight into the biology of risk and recovery. The sub-study included samples collected prior to treatment on Day 1 and at Day 8 from 303 patients. Among these 303 patients, 6/152 (3.9%) participants were hospitalized in the placebo group and 2/151 (1.3%) participants were hospitalized in the sotrovimab group.

We visualized the transcriptomes of each patient using Uniform Manifold Approximation and Projection (UMAP). We noted that from Day 1 to Day 8, the distribution of all transcriptome profiles tended to shift towards higher values of UMAP component 2 (**Figure 2a)**. We defined a putative risk cluster based on the differences in the distributions of Day 1 and Day 8 samples in the UMAP (see Methods). The described risk cluster includes Day 1 and Day 8 transcriptomics profiles for 6 of 8 hospitalized participants (**Figure 2b)**. Participants in the high-risk cluster were significantly older, white, with a higher NLR, and higher viral RNA levels in nasopharyngeal samples (**Suppl. Table S1**). The cluster analysis also highlighted that baseline seropositive participants were less likely to be associated with the high-risk transcriptome cluster on Day 1 and no seropositive patient remained in the high-risk cluster by Day 8 (**Figure 2c**).

**Figure 2.**
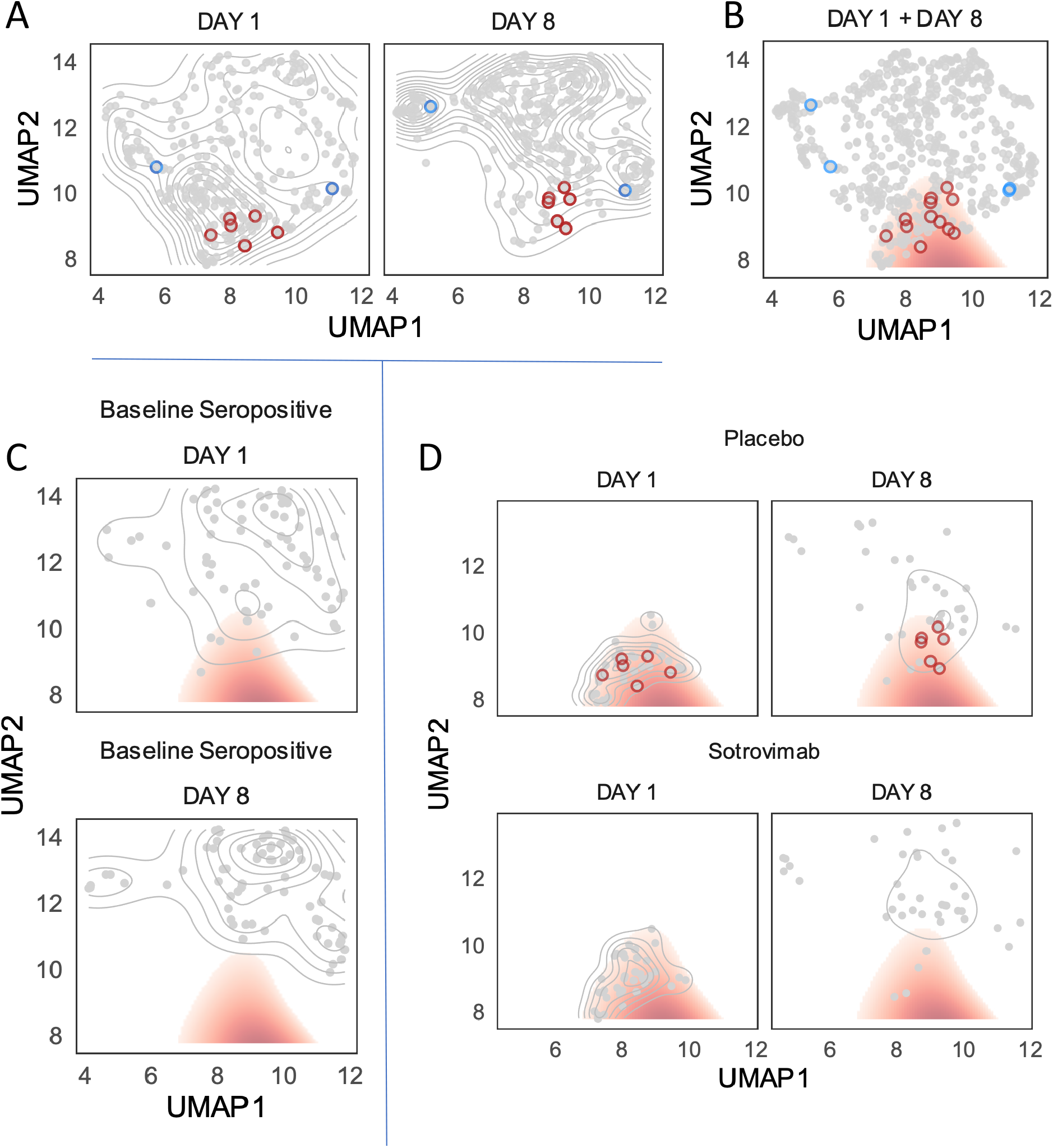
High-risk cluster defined by blood transcriptome profile. UMAP projection of transcriptomic profiles across Day 1 and Day 8 samples with hospitalized patients outlined in red (placebo) or blue (sotrovimab) circles. (**A**) A 2D kernel density, presented as a contour plot, highlight distribution of transcriptomics profiles in UMAP by visit day. (**B**) A threshold on the density difference between Day 1 and Day 8 distributions defines a high-risk cluster (red fill) which encompasses Day 1 and 8 transcriptomics profiles for 6 of 8 hospitalized patients. (**C**) Day 1 and Day 8 distributions of baseline seropositive patients (n=69). (**D**) Distribution of Day 1 and Day 8 transcriptomics profiles for patients in putative risk cluster at Day 1 split by treatment.

The two hospitalized participants mis-identified by the baseline transcriptome analysis were in the sotrovimab arm. One of the two participants had undetectable viral RNA in nasopharyngeal swabs at enrollment, and through 8 days post-enrollment when blood was drawn for the transcriptome analysis. This patient was then hospitalized by Day 21 with elevated viral load, suggestive of a unique clinical course. The second misidentified patient treated with sotrovimab was hospitalized due to a small intestinal obstruction deemed unrelated to COVID-19. Therefore, we found support for the hypothesis that the area outlined in red in **Figure 2** corresponded to an UMAP-defined high-risk cluster for COVID-19 progression where protective responses had failed to engage appropriately between Day 1 and Day 8. Although statistical power was limited due to only 8 hospitalizations in the transcriptomic sub-study, the transcriptome high risk group demonstrated a strong association to all-cause hospitalization and death (Fisher’s exact p=0.004) with a sensitivity of 75% [41%-94%] and a specificity of 76% [71%-80%].

### Response to treatment identified by transcriptomics

Given the effect of sotrovimab demonstrated in COMET-ICE, we determined whether treatment altered the probability of remaining in the transcriptome-defined high-risk cluster. To perform this analysis, we compared the rate of exiting the high-risk cluster for participants receiving sotrovimab versus placebo (**Figure 2d**). Among those who were high risk on Day 1, on Day 8, 29% of placebo-treated (n=11, including the hospitalized participants) versus 10% of sotrovimab-treated participants (n=4) remained high risk as defined by the transcriptome analysis. This corresponds to a 2.8-fold lower prevalence of risk-correlated transcriptional signatures for sotrovimab relative to placebo (Fisher’s exact p=0.045). Receipt of sotrovimab was also associated with a more rapid decline in viral RNA in nasopharyngeal samples by Day 8 (**Figure 3**).

**Figure 3.**
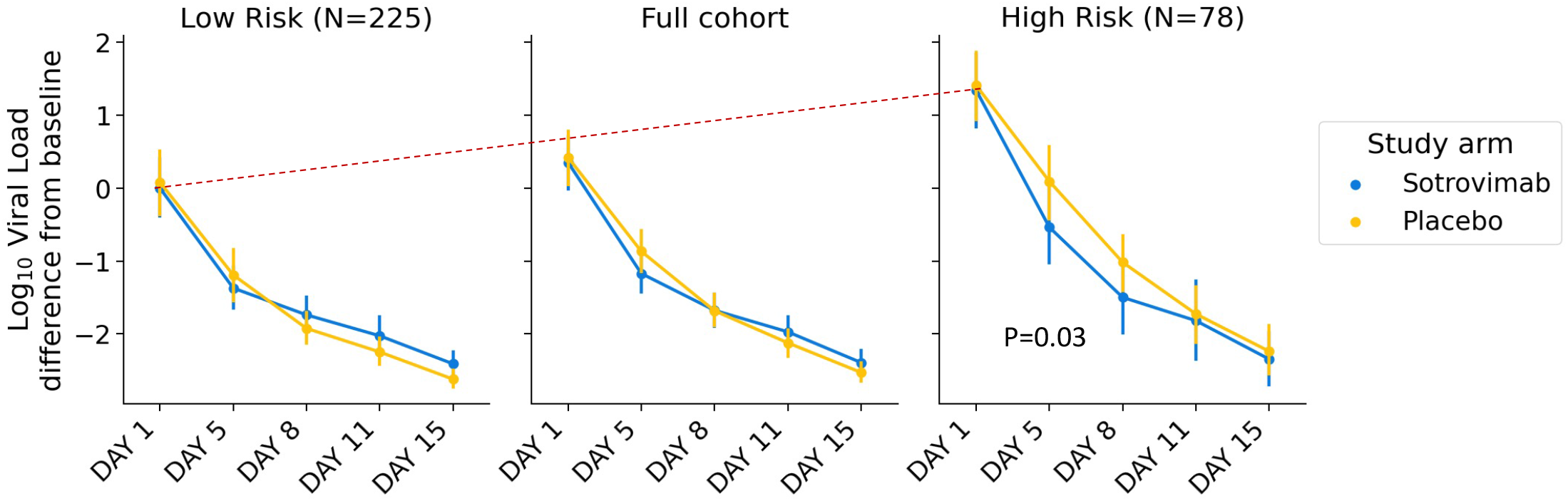
Viral RNA response to sotrovimab in high-risk transcriptome cluster. The high-risk transcriptome cluster associates with higher viral RNA concentration in respiratory secretions at both Day 1 and Day 8. The red dotted line highlights viral load differences at baseline between the groups. Error bars indicate the 95% confidence interval on the mean. At baseline, the high risk group had a viral load 1.1 log units higher than the cohort as a whole. At Day 5, the high risk cluster had a log viral load of 6.1 units in the placebo group, compared to 5.4 units in the sotrovimab group.

### Examining the biology of the transcriptome-defined high-risk cluster

For both Day 1 and Day 8 visits, we scored genes for differential expression between high versus low-risk clusters. We found a widespread transcriptional shift with thousands of genes identified as differentially expressed after adjusting for multiple comparisons (**Figure 4a, Suppl. File 2**). We characterized differentially expressed genes via gene-set enrichment analysis using the MSigDB Hallmark Gene Set annotation^14^. The most enriched Hallmark Gene Sets were associated with innate immune responses, in particular the complement system set, inflammatory response set, as well as the interferon alpha and gamma response gene expression modules (**Figure 4b**). Overexpression of genes in these pathways agrees well with previous work showing strong associations between increased innate immune system activation and disease severity^15^. In summary, whole transcriptome analysis is consistent with a significant inflammatory response and identifies participants on Day 1 that have a high risk of disease progression, a finding that is further supported by the lack of normalization of the high-risk transcriptome profile in participants who were subsequently hospitalized.

**Figure 4.**
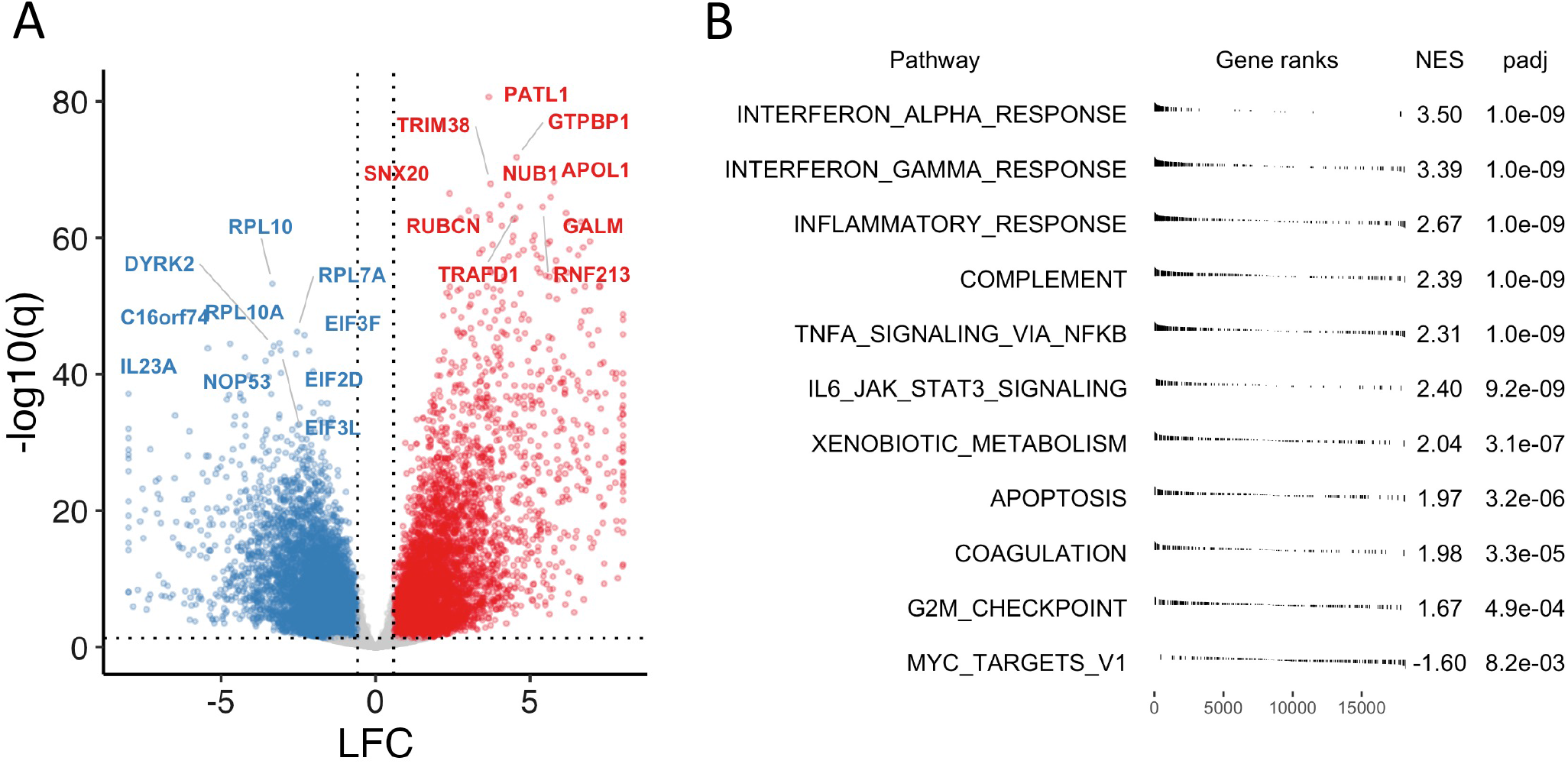
Transcriptome characteristics of high-risk group. (**A**) Summary of differential expression analysis results comparing high risk group to recovery group, accounting for visit day and subject gender, shown per gene with labels for top 10 among down-regulated (blue) and up-regulated (red) genes by statistical significance, respectively (q < 0.05, absolute LFC > log2(1.5)). For display, abs(LFC) <= 8. (**B**) Gene set enrichment analysis results using Hallmark Gene Sets (top 10 gene sets with q < 0 for NES > 0; q < 0.05 for NES < 0). LFC: log fold change. NES: Normalized enrichment score. q: FDR-adjusted p-value.

### Identifying a set of genes whose expression captures the risk-defining elements of the overall transcriptome

Having established a transcriptomic profile associated with risk of COVID-19 progression, recovery, and treatment response, we next determined whether a smaller number of mRNAs that might practically be measured by RT-PCR captured the predictive power of the overall transcriptome. Such an approach is preferable due to lower cost and greatly reduced turnaround time relative to whole transcriptome sequencing. To select a gene panel, we clustered genes into 10 groups according to their co-expression patterns across participants (**Suppl. Fig. S2**). This was accomplished using UMAP and K-means clustering. We then selected the top gene from each group as candidate for identification of a risk-predictive set of 10 genes. The 10 gene panel (*CD38, DAB2, EFHC2, EIF2D, EIF4B, MYO18A, NUDT3, OAS2, RPL10, TADA3*) accurately recapitulated the whole transcriptome risk clusters at both Day 1 (AUC=0.96) and Day 8 (AUC=0.99; **Figure 5a**). The expression of each gene in the panel is shown in **Figure 5b**. Expression of the 10-gene-panel was highly associated to viral load and hospitalization, and strongly affected by sotrovimab (**Suppl. Fig. S3**). On the transcriptomic subset, the 10-gene risk stratification had a sensitivity of 75% [41%-94%] and a specificity of 76% [72%-82%] for the prediction of all-cause hospitalization and death (Fisher’s exact p-value=0.003). The sensitivity increased to 83% and specificity to 80% when the analysis was limited to the placebo arm, reflecting the real case scenario where there is no modification of the outcome by sotrovimab. A comparison of this performance across risk predictors is presented in **Supp Table S2**.

**Figure 5.**
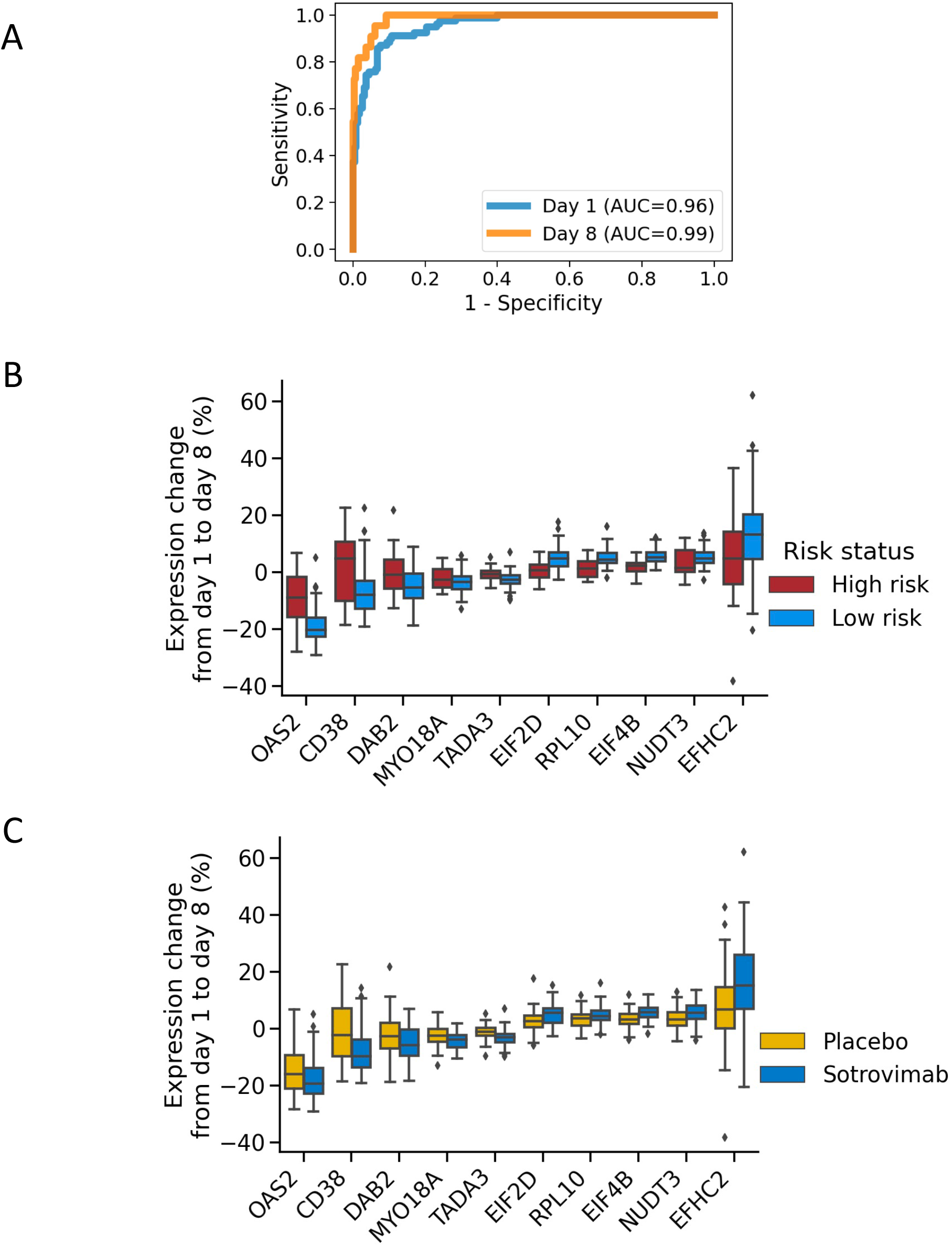
Surrogates to predict both risk of COVID-19 disease and response to sotrovimab using a 10 gene panel. (**A**) In cross-validation, the 10-gene panel accurately predicts risk groups assigned by the full transcriptome, at both day 1 and day 8. (**B**) Changes in expression for each of the genes in the 10-gene panel from day 1 to day 8. (**C**) Performance of each of the genes in the same 10-gene panel to track the response (change in expression from day 1 to day 8) to sotrovimab versus placebo.

## Discussion

We defined clinical laboratory and molecular biomarkers that can potentially identify participants with mild to moderate COVID-19 who are at highest risk of progression to severe disease and hospitalization or death using data collected in the prospective, phase 3 pivotal study COMET-ICE. Baseline NLR and a 10-gene transcriptomic signature associated with all-cause hospitalization or death with respective sensitivity and specificity of 33% and 93% (NLR) and of 87% and 64% (10-gene panel) on the transcriptomics sub-cohort. Changes in these biomarkers were also associated with response to treatment with the monoclonal antibody sotrovimab.

Currently, the risk of developing severe COVID-19 has been associated with a number of demographic factors such as age and specific comorbidities. However, there is considerable heterogeneity in disease outcome that would benefit from additional stratification of risk. NLR, the simple ratio of neutrophil over lymphocyte counts could be informative and easy to implement, an observation supported by other studies^16-18^ Though NLR sensitivity is low, the high specificity suggests that high NLR could be used as a triage test for persons at high risk of progressing and could prioritize those individuals for closer monitoring. Serostatus, defined here as IgG antibody response to nucleocapsid, is also a predictor of disease severity. None of the participants that were seropositive at baseline, whether because of previous infection or because of ongoing seroconversion^19^ died or were admitted to the ICU. Seropositive participants had lower levels of viral RNA in nasopharyngeal samples and were less likely to present or maintain a risk transcriptome profile. One caveat of this analysis is that VOCs that have decreased sensitivity to antibodies induced by vaccination (<u>e.g., Delta and Omicron)</u> were not circulating at the time of enrollment in the study. Therefore, additional analysis may be needed to confirm the protective effect of seropositivity in the current phase of the pandemic.

Whole blood transcriptome analysis revealed a signature of disease severity that encompassed overexpression of genes involved in interferon response, inflammation and the complement system. We showed that a full transcriptome signature can be captured faithfully with a 10-gene panel. Use of a simple expression signature lowers the bar for an eventual implementation, as recently shown by work to make a three gene tuberculosis signature using point-of-care rapid testing^20^.

An important effort of the present work was to define whether the set of predictive parameters of hospitalization and disease severity was also modified by treatment with sotrovimab; i.e., whether these parameters could serve as surrogate markers of sotrovimab response because they are modified by treatment and strongly associated to the study clinical endpoints of interest. Indeed, sotrovimab accelerated the normalization of NLR and the transcriptome profiles in a statistically significant manner. In particular, hospitalized participants in the placebo group retained the transcriptome profile associated with risk by Day 8 at a time when the majority of study participants normalized their peripheral blood gene expression profiles.

One topic of debate in the field is the value of measuring the levels of viral RNA in nasopharyngeal samples. In the present study, viral RNA levels measured by RT-PCR were of modest value as a baseline predictor. However, there was an association between viral RNA levels and the predictors of risk that we explored: NLR, serology and transcriptome profiles. Participants at risk of severe disease and hospitalization present higher levels of baseline nasopharyngeal viral RNA^21^. Weinreich et al.^22^ reported that mAb therapy had a significant effect on participants with a high viral load at baseline. Chen et al.^23^ reported a decreased viral load at day 11 did not appear to be a clinically meaningful end point, since the viral load was substantially reduced from baseline for the majority of patients, including those in the placebo group, a finding that is consistent with the natural course of the disease. Gottlieb et al.^24^ reported that treatment with mAb combination therapy, but not monotherapy, resulted in a reduction in SARS-CoV-2 log viral load at day 11 in participants with mild to moderate COVID-19. In the present work, resolution of disease was associated with decrease of viral load, in particular for participants receiving sotrovimab. However, taken together, these data indicate that viral load decline in upper airways is not a strong surrogate for clinical efficacy.

The strengths of the current study reside on the well-characterized, geographically diverse prospective pivotal clinical trial participant cohort with whole blood/RNA collected at multiple time points to evaluate response to treatment and disease characteristics over time. This dataset enabled the implementation of machine learning methods for predicting disease severity from both clinical and transcriptomic markers. A limitation of the study is that it included a pre-defined ‘risk population” of adults based on demographic and comorbid factors. A second limitation is the low number of study endpoints in the transcriptomic sub-study. However, if validated in additional studies, this approach could expand the definition of risk to include participants that might otherwise not be considered for treatment based on risk criteria currently in use^25^. In conclusion, this study identifies laboratory parameters associated with COVID-19 disease progression and hospitalization and shows that sotrovimab treatment effectively contributes to normalization of these parameters.

## Online Methods

### Characteristics of clinical trial population

COMET-ICE^2^ included 1057 adults with a positive local polymerase-chain-reaction or antigen SARS-CoV-2 test result and onset of symptoms within the prior 5 days (**Table 1**). Recruitment was between August 2020 and March 2021. Screening occurred within 24 hours prior to drug administration. Participants were required to be at risk for COVID-19 progression based on previously identified clinical parameters: age ≥55 years or adults with at least one of the following comorbid conditions: diabetes requiring medication, obesity (body-mass index >30 kg/m^2^), chronic kidney disease (estimated glomerular filtration rate <60 mL/min/1.73 m2), congestive heart failure (New York Heart Association class II or higher), chronic obstructive pulmonary disease, or moderate to severe asthma. Participants with already severe COVID-19, defined by shortness of breath at rest, oxygen saturation less than 94%, or requiring supplemental oxygen, were excluded. Participants were randomized 1:1 to receive either a single 500-mg infusion of sotrovimab or equal volume saline placebo. A subset of participants (n=303) consented for peripheral whole blood transcriptome analysis. Participants who opted-in to the transcriptome sub-study had similar demographic, clinical and laboratory characteristics to those in the overall study. They were evenly divided between placebo and sotrovimab arms (**Table 1**). In-person study visits occurred on days 1, 5, 8, 11, 15, 22 (W3), and 29 (W4) to assess adverse events and worsening of COVID-19. During study visits, blood samples and nasopharygeal swabs were collected for routine laboratory assessments and viral load, respectively. Samples for transcriptome analysis were collected twice: at the time of treatment (referred to as Day 1 herein) and a week later at the Day 8 visit. There was no analysis plan for this work in COMET-ICE; this was post-hoc analysis of the trial data.

### Clinical data analysis

The associations between laboratory values, and treatment response and hospitalization were measured using the area under a receiver operating characteristic curve (AUC). For single variable analyses, this metric was computed by directly ranking participants with no model fitting step, to avoid overfitting. Significance of AUC was assessed by the Mann-Whitney U test, relying on the equivalence between the Mann-Whitney U statistic and AUC. All reported AUCs were significant after a Bonferroni adjustment for multiple comparisons. The significance threshold was calculated as 0.05 divided by the number of clinical variables tested. For binary variables such as baseline risk factors, significance was assessed by Fisher’s exact test. Assessing complementarity of features was complicated by varying missingness patterns, leading to sample size loss. This was minimized by looking at only pairs of variables, and by median imputation of missing values^26^. Neither approach significantly improved on the single most predictive variable, indicating that single variable predictors are sufficient for accurate risk prediction.

### SARS-CoV-2 viral load and serology

Nasopharyngeal swabs were collected in universal transport media, and viral load was measured using the CDC 2019-nCoV Real-Time RT-PCR method run at central lab (https://www.fda.gov/media/134922/download). Serum samples were analyzed for anti-SARS-CoV-2 antibody by the Abbott SARS-CoV-2 IgG assay run on the Architect i2000SR immunoassay analyzer (https://www.fda.gov/media/137383/download). This assay qualitatively measures IgG anti-SARS-CoV-2 antibodies against the nucleocapsid protein. Due to the potential for cross-reaction of sotrovimab with anti-spike antibody assays, only analysis of anti-nucleocapsid serostatus was conducted.

### RNA isolation and sequencing

Peripheral whole blood was collected into PAXgene Blood RNA tubes (PreAnalytiX), identified by a sample accession number, and stored according to manufacturer recommendations. Day 1 and Day 8 samples for the same patient were sent for processing in the same batch. RNA purification, library preparation and sequencing were performed by Q2 Solutions – EA Genomics (Morrisville, NC). Total RNA was isolated (>1.25 ug required) and was depleted of globin mRNA using the GLOBINclear kit (Invitrogen). RNA quantity and quality was assessed using an Agilent Bioanalyzer (RIN score > 7.0 required). The globin-depleted RNA was used to generate a sequencing library using the TruSeq stranded mRNA method (Illumina). Briefly, poly(T) oligonucleotides are used to select poly-adenylated RNAs from the total RNA after globin mRNA reduction which are then fragmented and converted to cDNA using random primers in two steps to maintain strand specific information. Libraries were sequenced on Illumina NovaSeq 6000 (multiple sequencing runs, multiple instruments) to a target sequencing depth of 25 million paired-end reads per sample at a minimum read length of 50 bp. Samples were automatically selected by Q2 Solutions for repeat library preparation and sequencing based on pre-defined quality control metrics for ribosomal RNA fraction (>10% rRNA aligned reads threshold for repeat).

### RNA-seq analysis

701 sequenced libraries from 638 whole blood samples were delivered from Q2 solutions by processing batch and sequencing run. Reads were cropped to a common read length of 50 bp and low-quality bases and adapters were further trimmed using Trimmomatic (v. 0.39)^27^. Trimmed reads less than 31bp were discarded. Trimmed sequenced reads per library were then aligned to a custom reference genome using STAR (v 2.7.3a)^28^ and to a custom reference transcriptome using Salmon (v. 1.0.0)^29^. The custom reference genome and transcriptome annotation was based on combining the human reference genome and annotation from Gencode (GRCh38, release 30) with the SARS-CoV-2 reference genome and annotation from Ensembl (ASM985889v3 version). Libraries were assessed for total reads (minimum, maximum, and median of 23.4, 94.4, and 33.2 million read pairs), average read length (49 bp), and adapter content, post-trimming with FASTQC (v. 0.11.8). Alignment metrics, such as total aligned reads, aligned reads by feature type, gene body coverage and 3’ bias, were assessed using Picard CollectRnaSeqMetrics (v 2.20.2—0). We profiled duplication rate versus reads per kbp and verified that low read counts were not associated with high duplication at the library level using DupRadar (v 1.12.1). Known junction saturation rate as a function of sequencing depth was also profiled for all libraries using RSeQC (v 3.0.1). Transcript-level counts from Salmon were converted to gene-level counts and gene-level transcripts-per-million (TPM) using the R package tximport (v. 1.20.0). As genes with consistently low supporting read counts across libraries are unlikely to be called differentially expressed, a filtering step to remove genes with few to no supporting read counts across libraries was performed^30^. Conservatively, only genes with a minimum of 10 read counts in at least 4% (n=24) of the libraries were considered for further analysis (n = 23,540 genes). When multiple libraries (due to repeated library preparation and sequencing) were available for the same sample accession number (whole blood sample), a representative library with the higher median TPM value was selected as libraries with outlier values for alignment quality metrics were associated with (low) outlier median TPM values. Only libraries representing matched Day 1 and Day 8 whole blood samples for a patient were included for downstream analysis. No other library exclusion criteria were applied.

### Data analysis transcriptome

Using the R package DESeq2 (v 1.32.0)^31^, variance-stabilizing transformation was applied to gene-level counts.^30^ For exploration of transcriptome signatures, UMAP was run on the variance-stabilizing transformed RNA-seq count data. Prior to UMAP projection, data were pre-conditioned and de-noised using principal component a analysis (PCA). The first 20 PCs were selected based on the point at which explained variance tended towards zero (**Suppl. Fig. S4**). For this baseline analysis, UMAP (from the umap-learn python package; https://umap-learn.readthedocs.io/en/latest/) was run with default parameters.

To test the robustness of the embedding, this analysis was repeated for the full transcriptome (without PCA) for pathogen-associated transfer genes identified by di Iulio et al.^32^, and for immune-related pathway gene sets (Hallmark Gene Sets annotation).^14^ In each case, a variety of nearest neighbor values were tried, and the embedding was run multiple times to ensure repeatability. In all cases, the embeddings were similar. For example, the observed gradient between Day 1 and Day 8 samples, as well as the relative placement of the hospitalized was always consistent.

High risk due to laboratory parameters vs low risk categorizations were derived as follows. Two-dimensional kernel density estimation with a bandwidth of 1 was applied to Day 1 and Day 8 UMAP values separately. High risk participants were defined as those within an area where the Day 1 density exceeded the Day 8 density by 0.005. This cutoff was derived by choosing a round positive number near the beginning of the tail of the distribution (**Suppl. Fig. S5**). As a validation of this approach, we also performed a line search on this cutoff to optimize the separation between Day 1 and Day 8, as measured by Fisher’s exact p-value. This yielded an optimal cutoff of 0.006. To be conservative, we did not use this optimized value since the selection of cutoffs for the line search could be influenced by information beyond Day 1 vs Day 8 status.

Differentially expressed genes associated with the putative high-risk cluster were scored using a model accounting for gender and visit day (DESeq2, v. 1.32.0)^31^. Differentially expressed genes were characterized via gene set enrichment analysis (fgsea, v 1.18.0)^33^ using the Hallmark Gene Sets annotation (msigdbr, v. 7.4.1)^14^. For selection of a gene signature, we conducted diversity-based selection according to top ANOVA F-scores within 10 empirically identified gene clusters. Gene clusters were derived by performing UMAP dimensionality reduction on the transpose of the transcriptome matrix (with genes as rows instead of patients). This created a similarity map of genes based on their co-expression patterns across patients. We then defined 10 gene clusters from this map using K-means clustering (**Suppl. Fig. S2)**. From each cluster, we selected the gene most associated with risk according to its ANOVA F-score. Diversity-based selection using gene clustering significantly improved on greedy selection based on F-score alone (**Suppl. Fig. S6a**) and yielded comparable performance to transfer-learned^32^ and hallmark gene sets (**Suppl. Fig. S6b**). To assess performance of this set of 10 genes representing co-expression clusters, we repeated this entire process within a five-fold cross-validation loop, including gene clustering. In this procedure the dataset is partitioned into five folds. For each of the five folds, we trained a model on the other four chunks to predict its values in an unbiased manner. To avoid overfitting due to patient-specific attributes, samples from the same patient on different days were always kept in the same fold.

## Data Availability

Data availability is subject to the clinical trial charter

## Supplementary materials

**Supplemental Table S1.** Comparison of representative baseline values between high risk and low risk patients according to transcriptome data. Numbers in parentheses represent the percent of all patients in that category when denoted with “%”; otherwise, they represent the standard deviation. The numbers outside the parentheses are counts when represented with “%” in the parentheses. Otherwise, they are the mean value of that variable. P-value significance thresholds are based on a Bonferroni correction.

|  |  | **High risk cluster**<br>**(n=116)** | **Low risk cluster**<br>**(n=188)** | **p-value** |
| --- | --- | --- | --- | --- |
| **Demographics** | Age | 60.4 (30.0-95.0) | 50.5 (19.0-97.0) | 9x10<sup>-8</sup> |
|  | Female gender | 60 (52%) | 110 (59%) | 0.28 (N.S.) |
| **Risk factors** | Age>=65 | 44 (38%) | 32 (17%) | 7x10<sup>-5</sup> |
|  | COPD | 6 (5%) | 10 (5%) | 1.00 (N.S.) |
|  | Chronic kidney disease | 3 (3%) | 6 (3%) | 1.00 (N.S.) |
|  | Congestive heart failure | 2 (2%) | 4 (2%) | 1.00 (N.S.) |
|  | Diabetes mellitus | 32 (28%) | 37 (20%) | 0.12 (N.S.) |
|  | Moderate to severe asthma | 26 (22%) | 53 (28%) | 0.28 (N.S.) |
|  | Obesity (bmi > 30) | 54 (47%) | 129 (69%) | 2x10<sup>-4</sup> |
| **Race/ethnicity** | Asian | 8 (7%) | 10 (5%) | 0.62 (N.S.) |
|  | Black or African American | 4 (3%) | 23 (12%) | 0.01 (N.S.) |
|  | Latino | 46 (40%) | 107 (57%) | 0.005 (N.S.) |
|  | Native American | 1 (<1%) | 0 (0%) | 0.38 (N.S.) |
|  | White | 57 (49%) | 48 (26%) | 4x10<sup>-5</sup> |
| **Clinical** | Log10 viral load | 7.5 (1.5) | 5.5 (2.2) | 5x10<sup>-13</sup> |
|  | Hemoglobin | 140.9 (14.3) | 135.9 (16.6) | 0.008 (N.S.) |
|  | Lymphocytes | 1.3 (0.553) | 1.9 (0.712) | 6x10<sup>-14</sup> |
|  | Neutrophils | 3.5 (1.6) | 3.9 (1.7) | 0.04 (N.S.) |
|  | NLR | 3.4 (2.3) | 2.3 (1.5) | 7x10<sup>-6</sup> |
|  | Platelets | 214.5 (61.6) | 254.4 (71.1) | 7x10<sup>-6</sup> |
|  | APTT | 26.3 (2.9) | 25.6 (3.0) | 0.009 (N.S.) |
|  | spO2_blood | 97.1 (1.7) | 97.6 (1.2) | 0.004 (N.S.) |
| **Endpoint** | Hospitalization or death | 6 (5%) | 2 (1%) | 0.05 (N.S.) |

**Supplemental Table S2.** Sensitivity and specificity for different approaches for defining a high-risk group. Hosp, hospitalization in the full trial. Hosp_placebo, hospitalization in placebo arm. NLR, neutrophil lymphocyte ratio. Tx, transcriptome. Sens, sensitivity. Spec, specificity. Eqn, equation. Ppv, positive predictive value. Npv, negative predictive value. P, p value.

|  | NLR>6 |  | NLR>6, tx cohort |  | Seronegative |  | TxHighRisk |  | TxHighRisk_10gene |  |
| --- | --- | --- | --- | --- | --- | --- | --- | --- | --- | --- |
|  | HospOrDe<br>ath | HospOrDe<br>ath_place<br>bo | HospOrDe<br>ath | HospOrDe<br>ath_place<br>bo | HospOrDe<br>ath | HospOrDe<br>ath_place<br>bo | HospOrDe<br>ath | HospOrDe<br>ath_place<br>bo | HospOrDe<br>ath | HospOrDe<br>ath_place<br>bo |
| **sensitivity** | 44.8%<br>[27.9%-<br>62.7%] | 45.8%<br>[27.3%-<br>65.3%] | 33.3%<br>[7.7%-<br>71.4%] | 25.0%<br>[2.8%-<br>71.6%] | 83.3%<br>[68.8%-<br>92.7%] | 86.7%<br>[71.3%-<br>95.3%] | 75.0%<br>[40.8%-<br>94.4%] | 100.0%<br>[67.0%-<br>100.0%] | 75.0%<br>[40.8%-<br>94.4%] | 83.3%<br>[44.2%-<br>98.1%] |
| **sens_eqn** | 13/29 | 11/24 | 2/6 | 1/4 | 30/36 | 26/30 | 6/8 | 6/6 | 6/8 | 5/6 |
| **specificity** | 94.7%<br>[93.0%-<br>96.0%] | 94.8%<br>[92.4%-<br>96.6%] | 93.1%<br>[89.5%-<br>95.7%] | 93.8%<br>[88.7%-<br>97.0%] | 21.6%<br>[19.0%-<br>24.4%] | 20.9%<br>[17.3%-<br>24.9%] | 75.6%<br>[70.5%-<br>80.2%] | 78.1%<br>[70.9%-<br>84.2%] | 76.9%<br>[71.9%-<br>81.5%] | 79.5%<br>[72.4%-<br>85.4%] |
| **spec_eqn** | 832/879 | 402/424 | 243/261 | 122/130 | 196/906 | 92/440 | 223/295 | 114/146 | 227/295 | 116/146 |
| **ppv** | 21.7%<br>[12.7%-<br>33.3%] | 33.3%<br>[19.2%-<br>50.3%] | 10.0%<br>[2.1%-<br>28.4%] | 11.1%<br>[1.2%-<br>41.4%] | 4.1%<br>[2.8%-<br>5.7%] | 7.0%<br>[4.7%-<br>9.9%] | 7.7%<br>[3.3%-<br>15.2%] | 15.8%<br>[6.9%-<br>29.7%] | 8.1%<br>[3.5%-<br>15.9%] | 14.3%<br>[5.7%-<br>28.5%] |
| **ppv_eqn** | 13/60 | 11/33 | 2/20 | 1/9 | 30/740 | 26/374 | 6/78 | 6/38 | 6/74 | 5/35 |
| **npv** | 98.1%<br>[97.0%-<br>98.9%] | 96.9%<br>[94.9%-<br>98.2%] | 98.4%<br>[96.2%-<br>99.5%] | 97.6%<br>[93.7%-<br>99.3%] | 97.0%<br>[94.0%-<br>98.8%] | 95.8%<br>[90.4%-<br>98.6%] | 99.1%<br>[97.2%-<br>99.8%] | 100.0%<br>[97.8%-<br>100.0%] | 99.1%<br>[97.2%-<br>99.8%] | 99.1%<br>[96.1%-<br>99.9%] |
| **npv_eqn** | 832/848 | 402/415 | 243/247 | 122/125 | 196/202 | 92/96 | 223/225 | 114/114 | 227/229 | 116/117 |
| **p** | 3.7e-09 | 8.0e-08 | 6.7e-02 | 2.5e-01 | 6.8e-01 | 4.8e-01 | 4.4e-03 | 1.8e-04 | 3.3e-03 | 2.6e-03 |

## Supplemental figures

**Suppl. Fig. S1.**
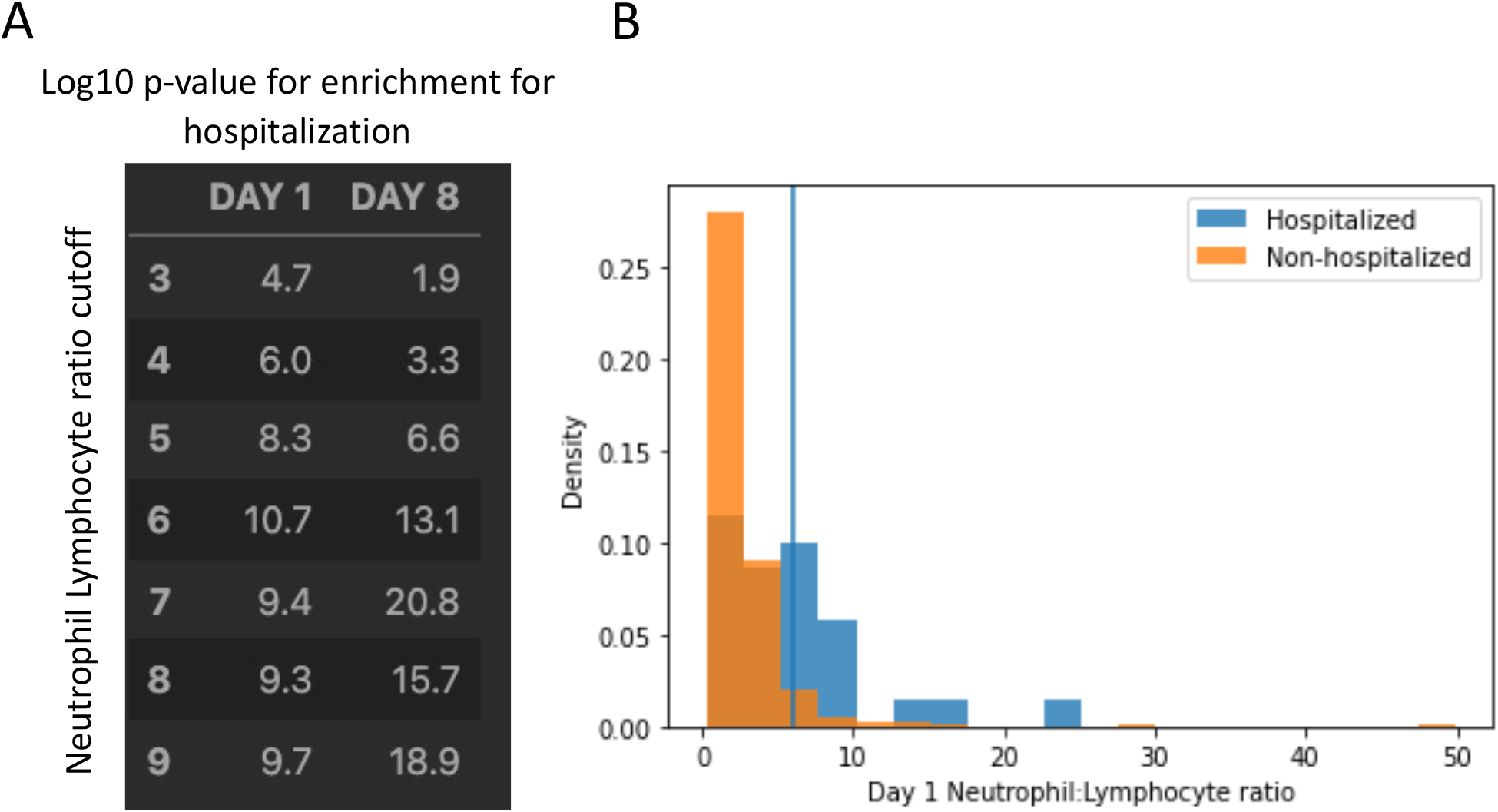
Threshold selection for neutrophil lymphocyte ratio. (**A**) Fisher’s exact p-value for association to hospitalization as a function of cutoff value and day. (**B**) a visualization of the distribution of Day 1 neutrophil lymphocyte ratio for hospitalized versus non-hospitalized patients

**Suppl. Fig. S2.**
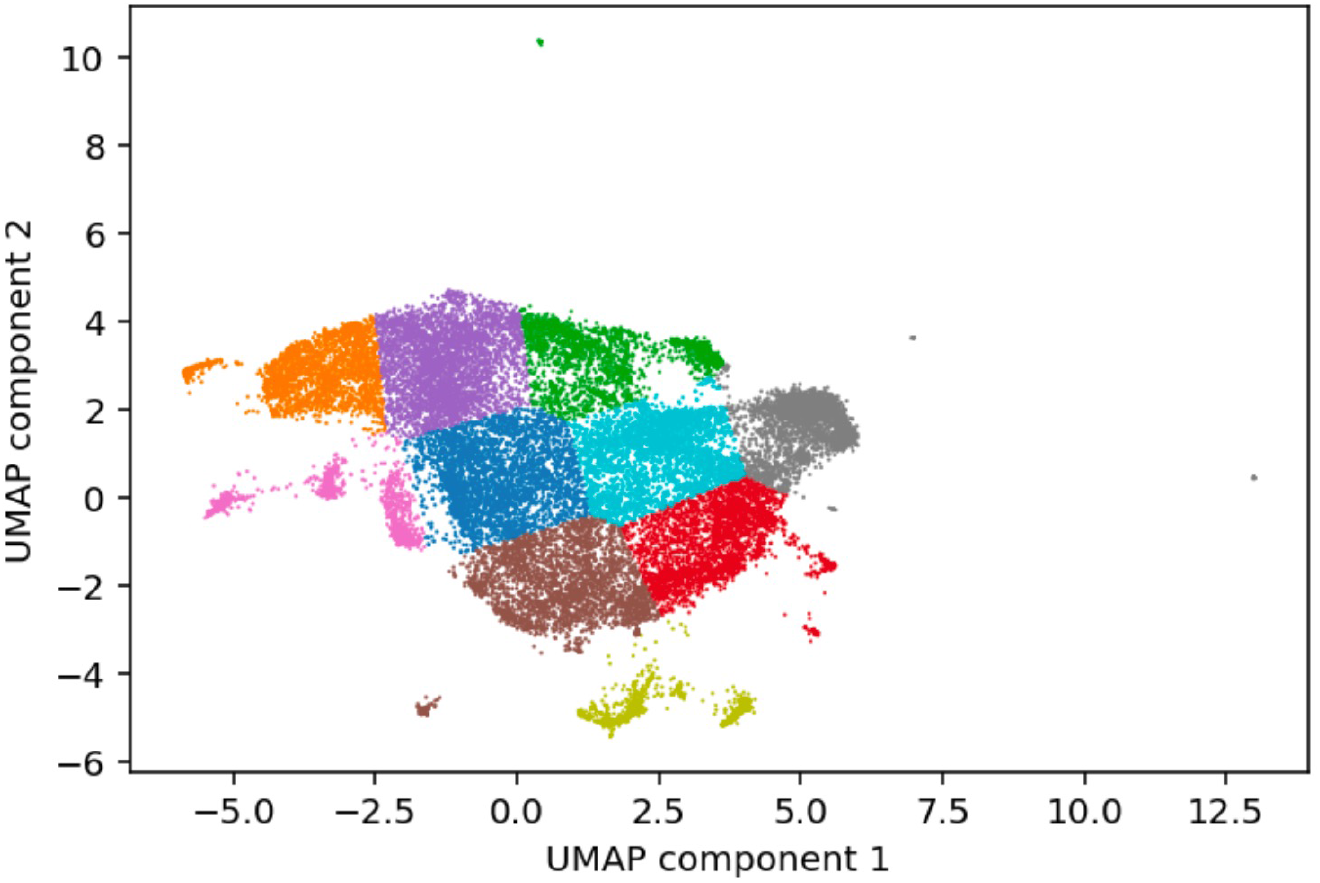
Grouping of genes based on their co-expression patterns across individuals. UMAP dimensionality reduction of genes based on transcriptomic patterns across patients. K-means clustering of genes into 10 groups.

**Suppl. Fig S3.**
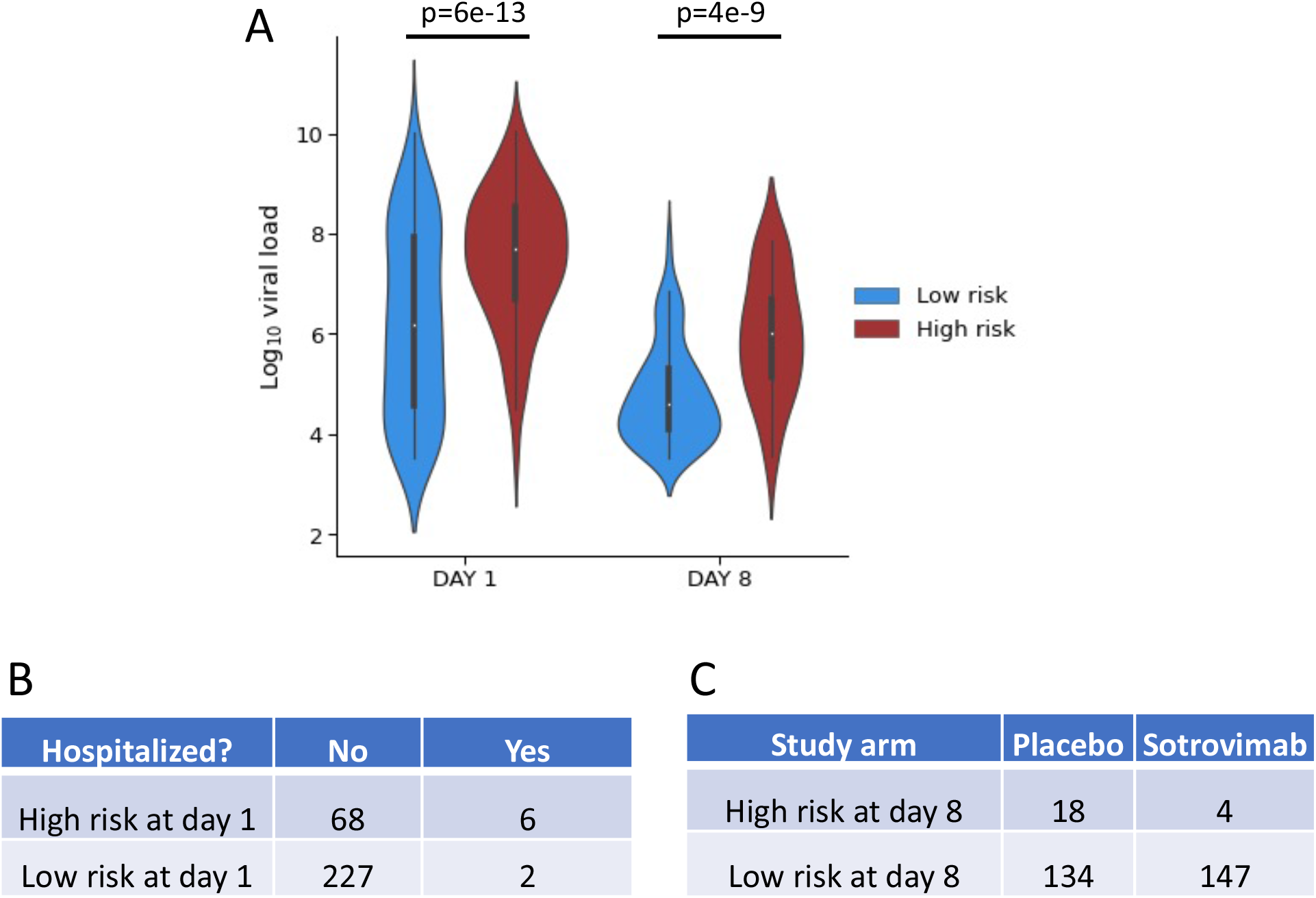
Surrogates of risk and recovery using a 10-gene panel. The 10-gene panel-derived risk groups separate patients by viral load at both day 1 and day 8. (**A**) The cross-validated, panel-derived risk assignments were associated to significantly higher viral load at both day 1 and day 8 (p<1e-5). (**B**) Day cluster status classified as high risk 6/8 samples from hospitalized patients (p=3e-3). (**C**) At day 8, 12% of patients (18 / 152) in the placebo group remained in the panel-derived risk group, compared to 3% in the sotrovimab group (4 / 151), corresponding to a risk reduction of 78% (p=3e-3).

**Suppl. Fig. S4.**
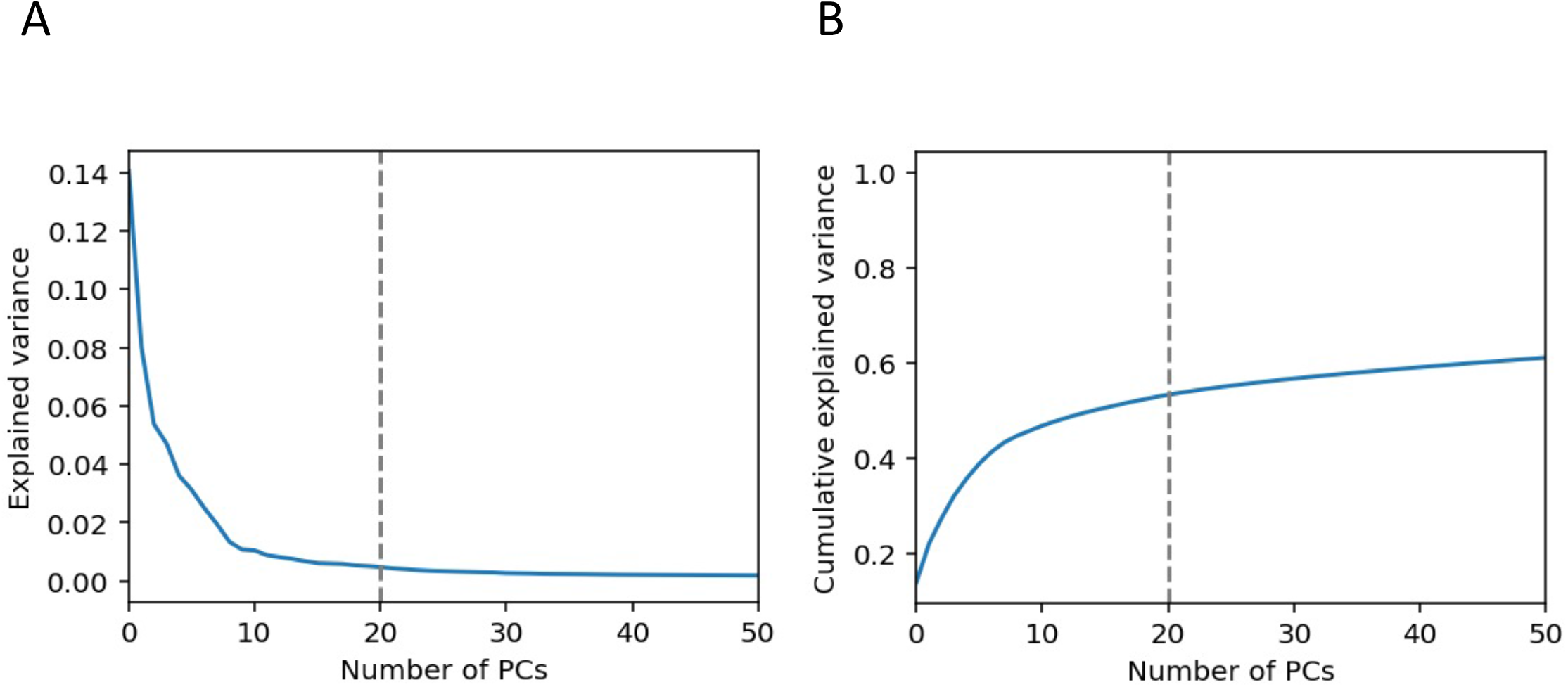
Selection of 20 PCs. (**A**) The explained variance of each principal component (**B**) the cumulative explained variance, summing from the first to Nth component, where N is denoted on the x-axis.

**Suppl. Fig. S5.**
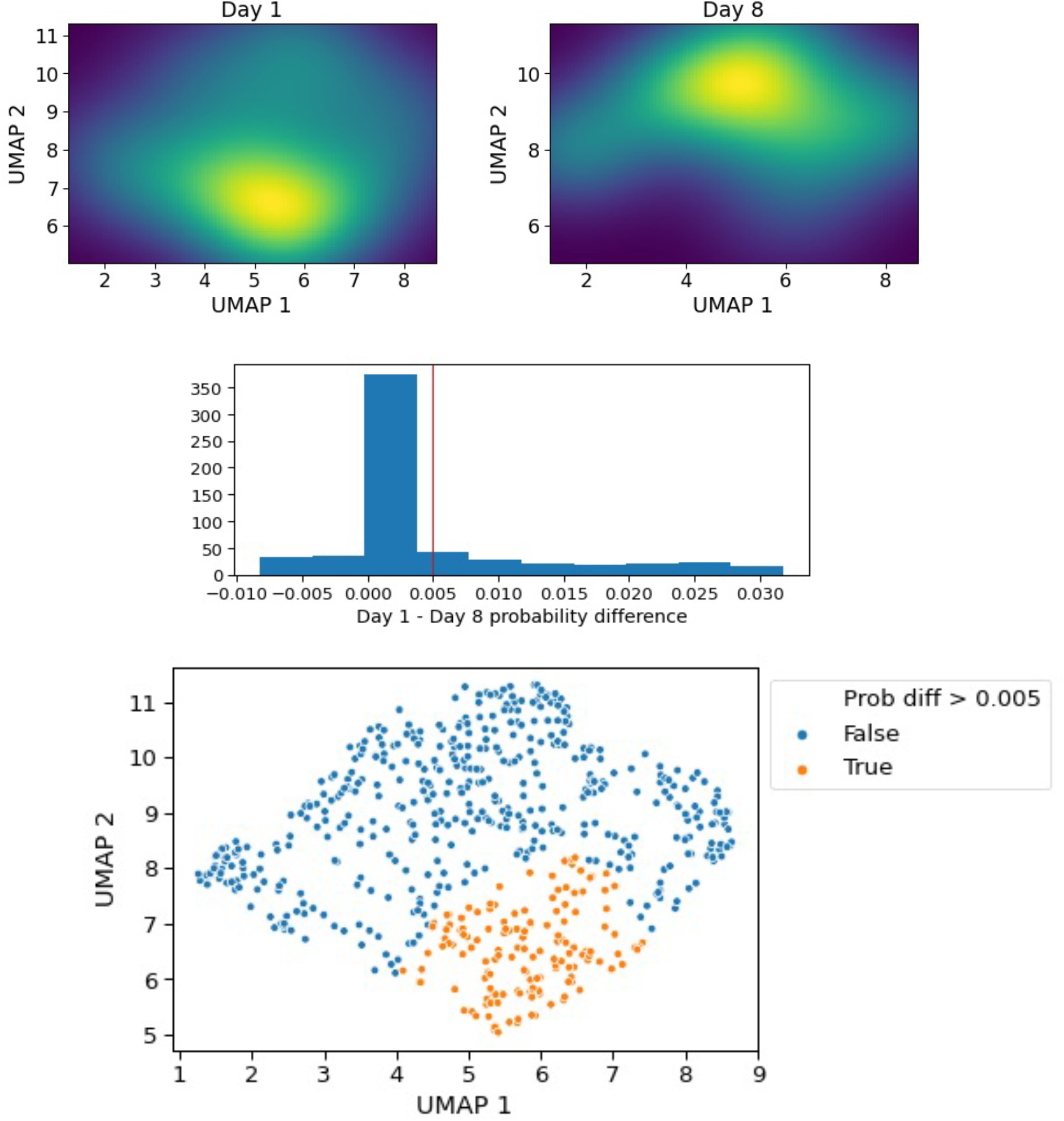
Definition of a high-risk cluster using transcriptome data. (**A**) kernel density estimators fit on Day 1 and Day 8 UMAP distributions. (**B**) The distribution of density differences between Day 1 and Day 8 for each patient. The red line denotes the chosen cutoff for defining a high-risk group. (**C**) A visualization of which patients are defined as high risk according to this cutoff.

**Suppl. Fig. S6.**
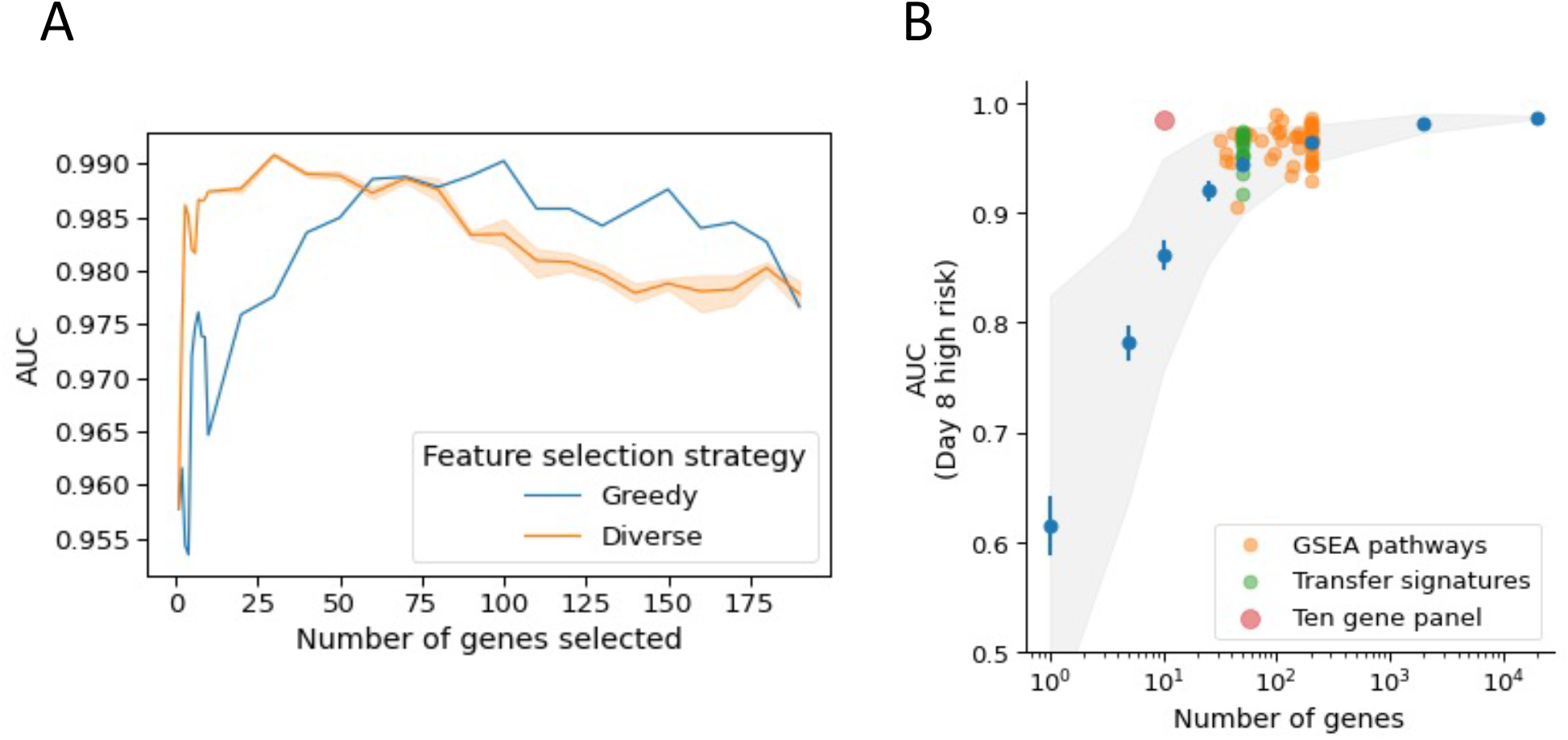
Predicting the high-risk cluster using a varying number of genes. **(A)** The blue line (AUROC) denotes selecting the top genes based on F-score. The orange line shows performance when the top F-scores are taken uniformly from each cluster. (**B**) Comparative performance of sources and number of genes of a predictive gene panel. AUROC as a function of the number of genes for predicting high risk group at day 8. Blue points and whiskers show the mean and confidence interval of the mean, respectively, for random gene sets. The grey envelope shows the 90% confidence interval for random gene performance across 100 replicates. Green points show the performance of the transfer signature gene sets published by di Iulio et al. Orange points demonstrate the performance of gene sets from GSEA pathways. Finally, the red point denotes the performance of the selected 10 gene panel.

